# Galantamine alleviates oxidative stress alongside anti-inflammatory and cardio-metabolic effects in subjects with the metabolic syndrome in a randomized trial

**DOI:** 10.1101/2019.12.30.19016105

**Authors:** Carine Teles Sangaleti, Keyla Yukari Katayama, Kátia De Angelis, Tércio Lemos de Moraes, Amanda Aparecida Araújo, Heno F. Lopes, Cleber Camacho, Luiz Aparecido Bortolotto, Lisete Compagno Michelini, Maria Cláudia Irigoyen, Douglas P. Barnaby, Kevin J. Tracey, Valentin A. Pavlov, Fernanda Marciano Consolim Colombo

## Abstract

**Background:** The metabolic syndrome (MetS) is an obesity-driven disorder with pandemic proportions and limited treatment options. Oxidative stress, low-grade inflammation and altered autonomic regulation, are important components of MetS pathophysiology. We recently reported that galantamine, an acetylcholinesterase inhibitor and an FDA-approved drug (for Alzheimer’s disease) alleviates the inflammatory state in MetS subjects. Here we examined the effects of galantamine on oxidative stress in parallel with inflammatory and cardio-metabolic parameters in subjects with MetS.

**Methods:** The effects of galantamine treatment, 8 mg daily for 4 weeks, followed by 16 mg daily for 8 weeks or placebo were studied in randomly assigned subjects with MetS (n=22 per group) of both genders. Oxidative stress, including superoxide dismutase (SOD), catalase (CAT), and glutathione peroxidase activities, lipid and protein peroxidation, and nitrite levels were analyzed before and at the end of the treatment. In addition, plasma cytokine and adipokine levels, insulin resistance (HOMA-IR) and other relevant cardio-metabolic indices were analyzed. Autonomic regulation was also examined by heart rate variability (HRV) before treatment, and at every 4 weeks of treatment.

**Results:** Galantamine treatment significantly increased antioxidant enzyme activities, including SOD (+1.65 USOD/mg protein, [95% CI 0.39 to 2.92], P=0.004) and CAT (+0.93 nmol/mg, [95% CI 0.34 to 1.51], *P*=0.011), decreased lipid peroxidation (thiobarbituric acid reactive substances, -5.45 pmol/mg, [95% CI -10.97 to 0.067], *P*=0.053) and systemic nitrite levels (-0.05 nit/mg protein, [95% CI -0.21 to 0.10], *P*=0.038) compared with placebo. In addition, galantamine significantly alleviated the inflammatory state and insulin resistance, and decreased the low frequency/high frequency ratio of HRV, following 8 and 12 weeks of drug treatment.

**Conclusion:** Low-dose galantamine alleviates oxidative stress, alongside beneficial anti-inflammatory, and metabolic effects, and modulates autonomic regulation in subjects with MetS. These findings are of considerable interest for further studies with galantamine to ameliorate MetS pathophysiology.

## Background

The metabolic syndrome (MetS), comprising a combination of central (abdominal) obesity, dyslipidemia, and elevated fasting glucose and blood pressure is a disorder of pandemic proportions. MetS is linked to a significantly increased risk for type 2 diabetes, cardiovascular disease, stroke, and other debilitating and life-threatening diseases (1, 2). Despite the enormous and growing deleterious impact of MetS on our modern societies and health care, finding treatments for this disorder as a whole has been challenging and remains inefficient. Low-grade chronic inflammation and increased oxidative stress have been identified as substantial contributing factors and drivers of MetS pathology (1, 3-5). In addition, autonomic nervous system dysfunction, with increased sympathetic and decreased vagus nerve activities, has been documented in MetS patients (6-10) and related to arterial hypertension, diabetes, heart failure, and stroke (11, 12). Accordingly, lowering chronic inflammation and oxidative stress, and improvement of autonomic nervous system function are promising therapeutic approaches in MetS (5, 8, 10, 13-15).

In addition to its “classical” physiological functions in autonomic regulation, the vagus nerve controls inflammation through a brain-integrated physiological mechanism termed the inflammatory reflex (16, 17). The inflammatory reflex is reportedly activated by administering centrally-acting cholinergic compounds, including galantamine - an acetylcholinesterase inhibitor and a clinically-approved drug for Alzheimer’s disease (17-25). Galantamine alleviates inflammation and metabolic derangements in murine models of many diseases, including high-fat diet-induced obesity and MetS (21, 26-29). In addition, galantamine alters autonomic nervous system modulation towards parasympathetic (vagus nerve) predominance (28, 30). Recently, in a randomized, double-blind placebo-controlled clinical trial we demonstrated that 12 weeks of administration of low to moderate galantamine doses (which are clinically approved), alleviated systemic inflammation and insulin resistance, and altered autonomic regulation in patients with MetS (31).

In addition to inflammation, oxidative stress is an important contributor to MetS mechanisms and a driver of pathology (5, 13). A prolonged state of oxidative stress is manifested by reduced activities of antioxidant enzymes and increased lipid peroxidation (3, 4, 32). The redox system imbalance may negatively affect the clustering components of MetS, and has been linked to atherogenesis (13, 32). Therefore, in this study we evaluated the effects of 12 weeks of galantamine treatment on oxidative stress, by analyzing superoxide dismutase (SOD), catalase (CAT) and glutathione peroxidase (GPx) activities, lipid and protein oxidation, and nitrate levels in these MetS patients. In parallel, we reexamined the anti-inflammatory, and other galantamine effects, and investigated the time course of galantamine autonomic modulation by assessing heart rate variability (HRV) components.

## Methods

### Study Design

The present study is based on analysis of data obtained from a prospective, randomized, double-blind, placebo-controlled trial with galantamine treatment in patients with the MetS. Results from this study, highlighting the drug effect on inflammation, HRV and cardio-metabolic parameters were recently published (31). The study protocol was reviewed and approved by the Institutional Ethical Committee and the Human Subject Protection Committee of Heart Institute (InCor) and Clinic Hospital of the University of São Paulo, Brazil (number 11672/555738), and registered at ClinicalTrials.gov (NCT02283242). It was conducted following the World Medical Association International Code of Medical Ethics. (Declaration of Helsinki, 1964; revised in 2008).

The complete study protocol was previously published (31). Briefly, participants in the study were of both genders, aged 18-59 years. They were diagnosed with MetS according to the ATP III criteria consisting of the presence of at least 3 of the following 5 parameters: increased abdominal circumference (≥102 cm for men and ≥ 88 cm for women); low plasma HDL cholesterol levels (< 40 mg/dl for men and <50 mg/dl for women); increased values for plasma triglycerides (≥ 150 mg/dl); elevated blood pressure (≥ 130 mmHg systolic blood pressure or ≥85 mm Hg diastolic blood pressure), and increased plasma glucose levels (≥100mg/dl). After the initial screening and consideration of inclusion and exclusion criteria, 60 eligible subjects of both genders were consecutively randomized in a 1:1 ratio to treatment with galantamine or placebo for 12 weeks. After recapsulation, galantamine hydrobromide extended-release capsules of 8 mg, commercially available as Reminyl® ER (manufactured by Janssen-Cilag Pharmaceuticals, Johnson & Johnson, Brazil), were administered in a dose of one tablet per day (8 mg/day) for four weeks and then titrated to 2 tablets per day (16 mg/day) for eight weeks. At each medical visit, the exact number of galantamine or placebo capsules were given to the patients. Pill counting in the next visit confirmed the adherence to the treatment. All patients with some exceptions previously noted (Ref) completed the study. In the course of the study, 44 (22 per arm) out of 60 patients had the Finometer determination of blood pressure, which was used for HRV analysis, recorded at all four key time points of the study. These were: T0 - basal measurements before treatment; T1 – following 4 weeks of galantamine 8 mg/day; T2 – following 4 weeks of galantamine 16 mg/day; and T3 - after 4 final weeks of galantamine 16 mg/day.

### Determination of oxidative and nitrosative stress, metabolic profile and inflammatory mediators

Venous blood samples were withdrawn twice, at baseline (T0) and the end of the protocol (T3). Parameters of oxidative stress were determined in plasma as previously described (11). Quantification of superoxide dismutase (SOD) activity was performed based on the inhibition of the reaction between O_2_ and pyrogallol (33). Catalase (CAT) activity was determined by measuring the reduction in H_2_O_2_ absorbance at 240nm (34, 35). Glutathione peroxidase (GPx) activity was based on the consumption of NADPH (22). For the TBARS assay (lipid peroxidation), trichloroacetic acid (10%, w/v) was added to the homogenate for proteins precipitation and sample acidification (36). The mixture was centrifuged (3000□g, 3□min), the protein-free sample extracted and thiobarbituric acid (0.67%, w/v) was then added to the reaction medium and tubes were placed in a water bath (100□°C) for 15 min. Absorbance was measured at 535□nm using a spectrophotometer (36). The protein oxidation was measured by a reaction of protein carbonyl groups with 2,4-dinitrophenylhydrazine to form 2,4-dinitrophenylhydrazone, which can be quantified spectrophotometrically. The reaction product was measured at 360 nm (36). The Griess reagent was used to determine the plasma nitrites (NO^−2^) (11). Blood tests, including triglycerides, total, HDL and LDL cholesterol as well as fasting glucose, were performed according to standard protocols. Plasma samples were stored at –80°C before analysis. Cytokines, adipokines and insulin were analyzed using multiplex immunoassay (all from Millipore): HCYTMAG-60K-PX41 for TNF; HADK2MAG-61K for leptin and insulin; and HADK1MAG-61K for adiponectin. Individual values for leptin and adiponectin were used to calculate the leptin/adiponectin ratio as previously described (31).

### Hemodynamic parameters and heart rate variability (HRV) evaluation

All subjects were asked to abstain from exercise for 24h prior to study visits and from drinking caffeinated products on the morning of their evaluation. During a 15 minute study period, subjects were supine and awake in a quiet room while blood pressure waveforms (BP) were captured and stored using a digital photoplethysmograph device (Finometer, Finapres Medical System BV, Holland) as previously described (31). Recordings were visually inspected to remove non-stationary data and sequential pulse intervals (PI) and then used to compute HRV in both frequency and time domains as previously described in detail (31). All analyses adhered to standards developed by the Task Force of the European Society of Cardiology and the North American Society of Pacing and Electrophysiology (37).

### Statistical analysis

Descriptive statistics were calculated separately for each of the two treatments (galantamine and placebo) using mean ± standard deviation and median (25^th^ percentile, 75^th^ percentile) for continuous data, and frequencies and percentages for categorical data. To establish baseline comparability of the two randomized treatment arms, the chi-square test was used to compare categorical variables (i.e., gender) and the Mann-Whitney test, the non-parametric counterpart to the two-sample t-test, was used to compare continuous measures (**Table 1**).

**Table 1.**
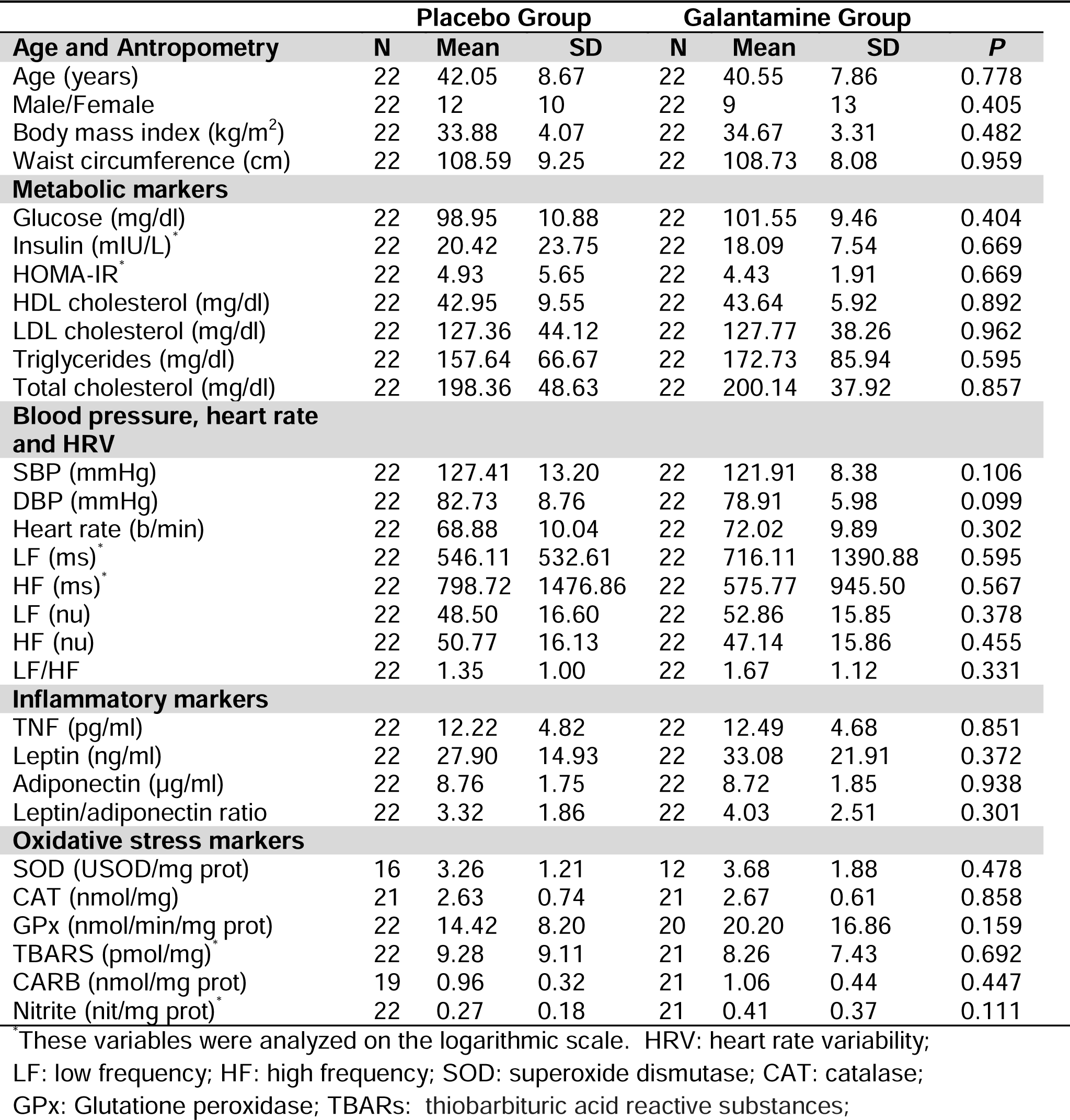
Baseline characteristics

|  | Placebo Group |  |  | Galantamine Group |  |  |  |
| --- | --- | --- | --- | --- | --- | --- | --- |
| **Age and Antropometry** | **N** | **Mean** | **SD** | **N** | **Mean** | **SD** | **P** |
| Age (years) | 22 | 42.05 | 8.67 | 22 | 40.55 | 7.86 | 0.778 |
| Male/Female | 22 | 12 | 10 | 22 | 9 | 13 | 0.405 |
| Body mass index (kg/m<sup>2</sup>) | 22 | 33.88 | 4.07 | 22 | 34.67 | 3.31 | 0.482 |
| Waist circumference (cm) | 22 | 108.59 | 9.25 | 22 | 108.73 | 8.08 | 0.959 |
| **Metabolic markers** |  |  |  |  |  |  |  |
| Glucose (mg/dl) | 22 | 98.95 | 10.88 | 22 | 101.55 | 9.46 | 0.404 |
| Insulin (mIU/L)* | 22 | 20.42 | 23.75 | 22 | 18.09 | 7.54 | 0.669 |
| HOMA-IR* | 22 | 4.93 | 5.65 | 22 | 4.43 | 1.91 | 0.669 |
| HDL cholesterol (mg/dl) | 22 | 42.95 | 9.55 | 22 | 43.64 | 5.92 | 0.892 |
| LDL cholesterol (mg/dl) | 22 | 127.36 | 44.12 | 22 | 127.77 | 38.26 | 0.962 |
| Triglycerides (mg/dl) | 22 | 157.64 | 66.67 | 22 | 172.73 | 85.94 | 0.595 |
| Total cholesterol (mg/dl) | 22 | 198.36 | 48.63 | 22 | 200.14 | 37.92 | 0.857 |
| **Blood pressure, heart rate and HRV** |  |  |  |  |  |  |  |
| SBP (mmHg) | 22 | 127.41 | 13.20 | 22 | 121.91 | 8.38 | 0.106 |
| DBP (mmHg) | 22 | 82.73 | 8.76 | 22 | 78.91 | 5.98 | 0.099 |
| Heart rate (b/min) | 22 | 68.88 | 10.04 | 22 | 72.02 | 9.89 | 0.302 |
| LF (ms)* | 22 | 546.11 | 532.61 | 22 | 716.11 | 1390.88 | 0.595 |
| HF (ms)* | 22 | 798.72 | 1476.86 | 22 | 575.77 | 945.50 | 0.567 |
| LF (nu) | 22 | 48.50 | 16.60 | 22 | 52.86 | 15.85 | 0.378 |
| HF (nu) | 22 | 50.77 | 16.13 | 22 | 47.14 | 15.86 | 0.455 |
| LF/HF | 22 | 1.35 | 1.00 | 22 | 1.67 | 1.12 | 0.331 |
| **Inflammatory markers** |  |  |  |  |  |  |  |
| TNF (pg/ml) | 22 | 12.22 | 4.82 | 22 | 12.49 | 4.68 | 0.851 |
| Leptin (ng/ml) | 22 | 27.90 | 14.93 | 22 | 33.08 | 21.91 | 0.372 |
| Adiponectin (µg/ml) | 22 | 8.76 | 1.75 | 22 | 8.72 | 1.85 | 0.938 |
| Leptin/adiponectin ratio | 22 | 3.32 | 1.86 | 22 | 4.03 | 2.51 | 0.301 |
| **Oxidative stress markers** |  |  |  |  |  |  |  |
| SOD (USOD/mg prot) | 16 | 3.26 | 1.21 | 12 | 3.68 | 1.88 | 0.478 |
| CAT (nmol/mg) | 21 | 2.63 | 0.74 | 21 | 2.67 | 0.61 | 0.858 |
| GPx (nmol/min/mg prot) | 22 | 14.42 | 8.20 | 20 | 20.20 | 16.86 | 0.159 |
| TBARS (pmol/mg)* | 22 | 9.28 | 9.11 | 21 | 8.26 | 7.43 | 0.692 |
| CARB (nmol/mg prot) | 19 | 0.96 | 0.32 | 21 | 1.06 | 0.44 | 0.447 |
| Nitrite (nit/mg prot)* | 22 | 0.27 | 0.18 | 21 | 0.41 | 0.37 | 0.111 |

\*These variables were analyzed on the logarithmic scale. HRV: heart rate variability;
LF: low frequency; HF: high frequency; SOD: superoxide dismutase; CAT: catalase;
GPx: Glutathione peroxidase; TBARS: thiobarbituric acid reactive substances;

Repeated measures analysis of variance (RMANOVA) with a mixed models approach was used to determine if the two groups behave differently over time (i.e., the group x time interaction) for each of the following measures: CAT, SOD, GPx, TBARS, carbonyls, nitrites, low frequency (LF) of HRV, high frequency (HF) of HRV (absolute and normalized values), LF/HF ratio, BMI, SBP, DBP, glucose, HDL cholesterol, LDL cholesterol, triglycerides, total cholesterol, TNF, leptin, adiponectin, and HOMA-IR. For all analyses, the standard assumptions of Gaussian residuals and equality of variance were tested. When normality assumption was not met, the logarithm transformation was used for the analysis of these variables. The repeated within-subjects factor was time (pre- and post), and the within-subjects factor was treatment group (galantamine and placebo). Data analyzed on the raw scale are reported as the arithmetic difference, calculated as post-minus pre-, and standard deviation (SD) for each group. “Positive” values indicate an increase in the measure from pre-to post- and “negative” values indicate decreases in the measure from pre- to post-. Those variables analyzed on the log scale were back transformed and summarized using the geometric mean ratio (GMR), which corresponds to the magnitude of difference between pre- and post-measurements; values >1 indicate an increase from pre-to post- and values <1 indicate a decrease from pre- to post-(**Tables 2 and 3**).

**Table 2.** Effect of galantamine on oxidative stress parameters

|  | Baseline |  |  |  |  |  | Follow-up |  |  |  |  |  | Treatment x Time Interaction |  |  |  |
| --- | --- | --- | --- | --- | --- | --- | --- | --- | --- | --- | --- | --- | --- | --- | --- | --- |
|  | Placebo Group |  |  | Galantamine Group |  |  | Placebo Group |  |  | Galantamine Group |  |  | Effect value | 95% (IC) |  | P |
| SOD (USOD/mg prot) | 16 | 3.26 | 1.21 | 12 | 3.68 | 1.88 | 16 | 2.80 | 21.17 | 12 | 4.45 | 2.06 | 1.65 | 0.39 | 2.96 | 0.004 |
| CAT (nmol/mg) | 21 | 2.63 | 0.74 | 21 | 2.67 | 0.61 | 21 | 3.06 | 0.82 | 21 | 3.99 | 1.05 | 0.93 | 0.34 | 1.51 | 0.011 |
| GPx (nmol/min/mg prot) | 22 | 14.42 | 8.20 | 20 | 20.20 | 16.86 | 22 | 17.96 | 14.12 | 20 | 18.56 | 14.05 | -1.64 | -9.31 | 6.03 | 0.329 |
| TBARS (pmol/mg) | 22 | 9.28 | 9.11 | 21 | 8.26 | 7.43 | 22 | 10.93 | 12.24 | 21 | 5.48 | 2.70 | -5.45 | -10.97 | 0.07 | 0.053 |
| CARB (nmol/mg prot) | 19 | 0.96 | 0.32 | 21 | 1.06 | 0.44 | 19 | 0.91 | 0.22 | 21 | 0.83 | 0.20 | -0.07 | -0.21 | 0.06 | 0.195 |
| Nitrite (nit/mg prot)* | 22 | 0.27 | 0.18 | 21 | 0.41 | 0.37 | 22 | 0.35 | 0.32 | 21 | 0.29 | 0.19 | -0.05 | -0.21 | 0.10 | 0.038 |

\*These variables were analyzed on the logarithmic scale. SOD: superoxide dismutase; CAT: catalase; GPx: glutathione peroxidase; TBARS: thiobarbituric acid reactive substances;

**Table 3.**
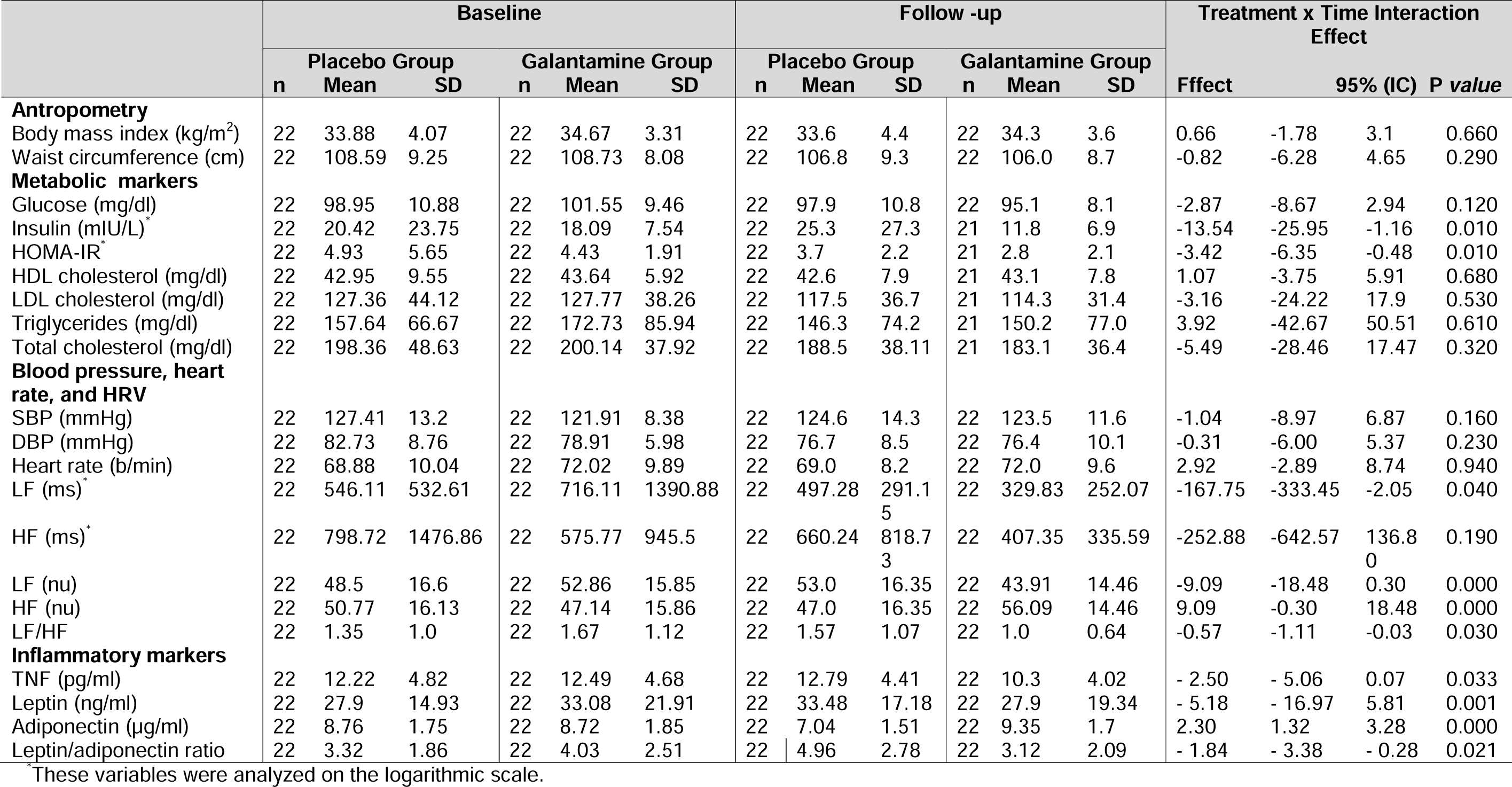
Effect of galantamine on metabolic, hemodynamic, and inflammatory markers.

The pairwise difference (pre-to post-) of the measurements within each treatment arm, as well as the difference in the placebo group and the galantamine group at the final time point, were determined using Tukey post-hoc analysis. Unless otherwise specified, a result was considered statistically significant at the p<0.05 level of significance. All analyses were performed using SPSS version 20.0 (SAS Institute Inc., Cary, NC).

## RESULTS

### Patients

Of the 60 patients enrolled in the original trial, 44 (22 in each arm) had complete HRV recordings available for analysis and were included in the current study (see **Figure 1**). Male: female ratios were 12/10 in the placebo group and 9/13 in the galantamine group. There were no statistically significant differences between the placebo and galantamine groups in the baseline values of all variables analyzed, including age and anthropometry, metabolic, hemodynamic, inflammatory, and oxidative stress indices (**Table 1**). It should be noted that some of the SOD values were below the lower detection limits of the assay and these values were omitted from the study. Therefore, the analysis of SOD levels were performed based on data from 16 subjects in the placebo group and 12 subjects in the galantamine group.

**Figure 1.**
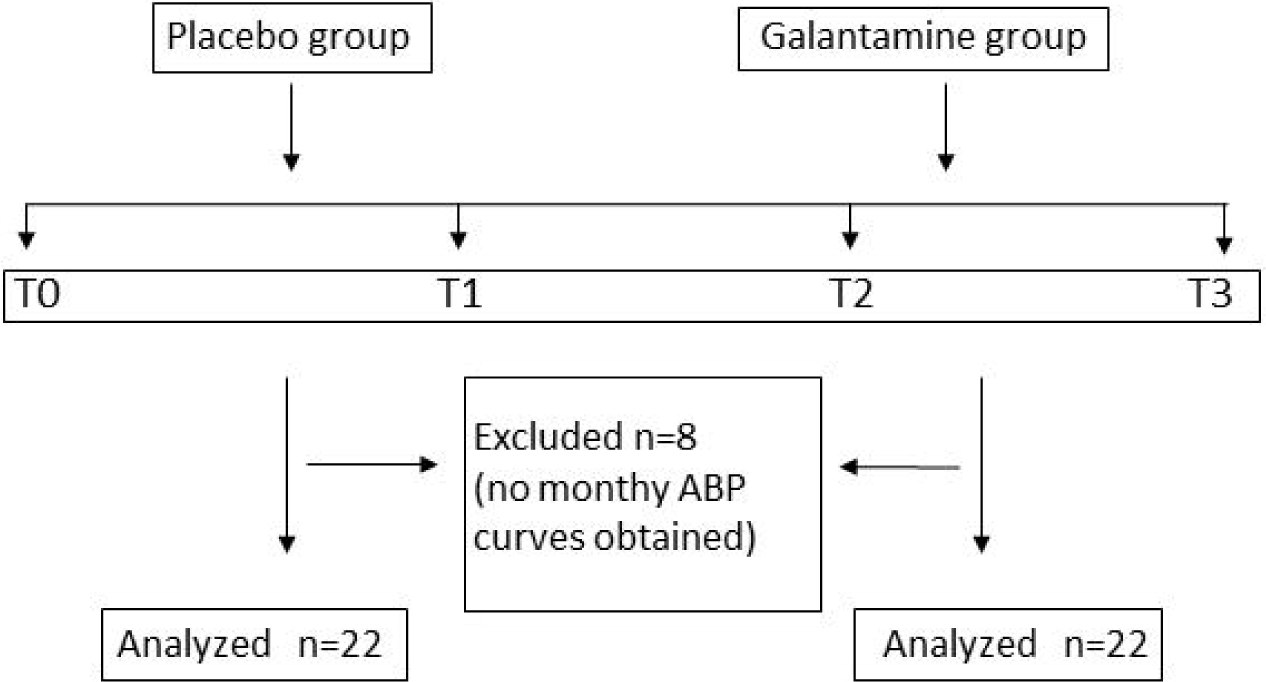
Flow chart (protocol sequence). T0 - blood sample draw, blood pressure curve recording; T1 and T2 – blood pressure curve recording; T3 – blood sample draw, blood pressure curve recording

### Effects of galantamine on oxidative and nitrosative stress indices

Administration of galantamine conferred significant protection against oxidative stress. At the end of the 12-week treatment period, the activities of antioxidant enzymes, including SOD and CAT increased significantly in the galantamine treated subjects compared with the placebo group (+1.65 USOD/mg protein, [95% CI 0.39 to 2.92], *P*=0.004) and (+0.93 nmol/mg, [95% CI 0.34 to 1.51], *P*=0.011), respectively (**Table 2**). No significant effect of galantamine treatment was detected on GPx activity (-1.64 nmol/min/mg prot, [95% CI -9.31 to 6.03], *P*=0.329) (**Table 2**). Galantamine decreased lipid peroxidation compared with placebo, as indicated by reduced levels of TBARS (a marker of oxidative damage (-5.45 pmol/mg, [95% CI -10.97 to 0.067], P=0.053) (**Table 2**). Galantamine treatment did not have a significant effect, compared with placebo, on protein peroxidation (determined by reaction of protein carbonyl groups) (-0.07 nmol/mg prot [95% CI -0.21 to 0.06], *P*=0.195) (**Table 2**). However, a significant reduction in systemic nitrite levels was observed in subjects treated with galantamine compared with those treated with placebo (-0.05 nit/mg prot, [95% CI -0,21 to 0,10], P=0,038) (**Table 2**).

### Effects of galantamine on inflammatory and metabolic indices

We reanalyzed plasma samples for inflammatory and metabolic indices to address the important question of whether galantamine effects on oxidative stress were associated with retaining beneficial anti-inflammatory and metabolic effects of galantamine (31) in the 22 patients per group. Adiponectin levels were significantly increased (+2,30 ug/ml [95% CI 1.32 to 3.28], P=0.0001) in the galantamine-treated patients compared with the placebo-treated (**Table 3**). In contrast, plasma TNF levels (-2.50 pg/ml [95% CI -5.06 to 0.07], P=0.033), and leptin levels (-5.18 ng/ml [95% CI -16.97 to 5.18], p=0.001), and leptin/adiponectin ratio (-1.84 [95% CI -3,38 to -0.28], P=0.021) were significantly decreased in patients treated with galantamine compared with the placebo-treated) (**Table 3**). In addition, galantamine treatment significantly decreased plasma insulin levels and HOMA-IR compared with placebo (P=0.01 for both effects) (**Table 3**). As previously reported (31) no significant differences between the groups were determined in BMI, waist circumference, HDL cholesterol levels, LDL cholesterol, triglyceride, and total cholesterol levels, systolic BP (SBP), diastolic BP (DBP), and heart rate (**Table 3**).

### Effects of galantamine on heart rate variability

We have previously reported that at the end of the 12-week treatment period there were significant changes in HRV parameters of MetS subjects treated with galantamine, compared with the placebo group, reflecting altered autonomic modulation (31). In this study, we found that these galantamine effects were preserved in the smaller cohorts (n=22) of MetS patients (**Table 3**). The low frequency (LF) power (ms^2^) of HRV in the galantamine group was significantly decreased, compared with the placebo group (log scale 0.48 ms^2^ [95% CI 0.29 to 0.80], *P =* 0.005) (**Table 3**). The LF/HF ratio was significantly decreased in the galantamine group compared with the placebo group (log scale 0.50 nu [95% CI 0.35 to 0.71], *P* <0.0001) reflecting the lower LF (normalized units [nu]) and the corresponding increase in high frequency (HF) (nu) (**Table 3**). Considering the importance of autonomic modulation in physiology and pathophysiological conditions (38), we further examined the time course of these drug effects on HRV in MetS subjects. Sequential analysis of HRV at T1 showed unaltered LF and HF (nu) after 4 weeks of galantamine (8 mg/day) treatment, compared with placebo (**Figure 2 A,B**). Increasing the drug dose to 16 mg/day for 4 weeks resulted in significantly higher HF and lower LF at T2 (P=0.007 and P=0.009, respectively) (**Figure 2 A,B**). These alterations were retained at T3 (end of study), after an additional 4 weeks of 16 mg/day galantamine treatment, compared with placebo (P=0.04 and P=0.005, respectively) (**Figure 2 A,B**). Accordingly, the LF/HF ratio, a proposed index of sympatho-vagal balance was significantly decreased at T2 (P=0.001) and T3 (P=0.001) compared with placebo (**Figure 2 C**).

**Figure 2.**
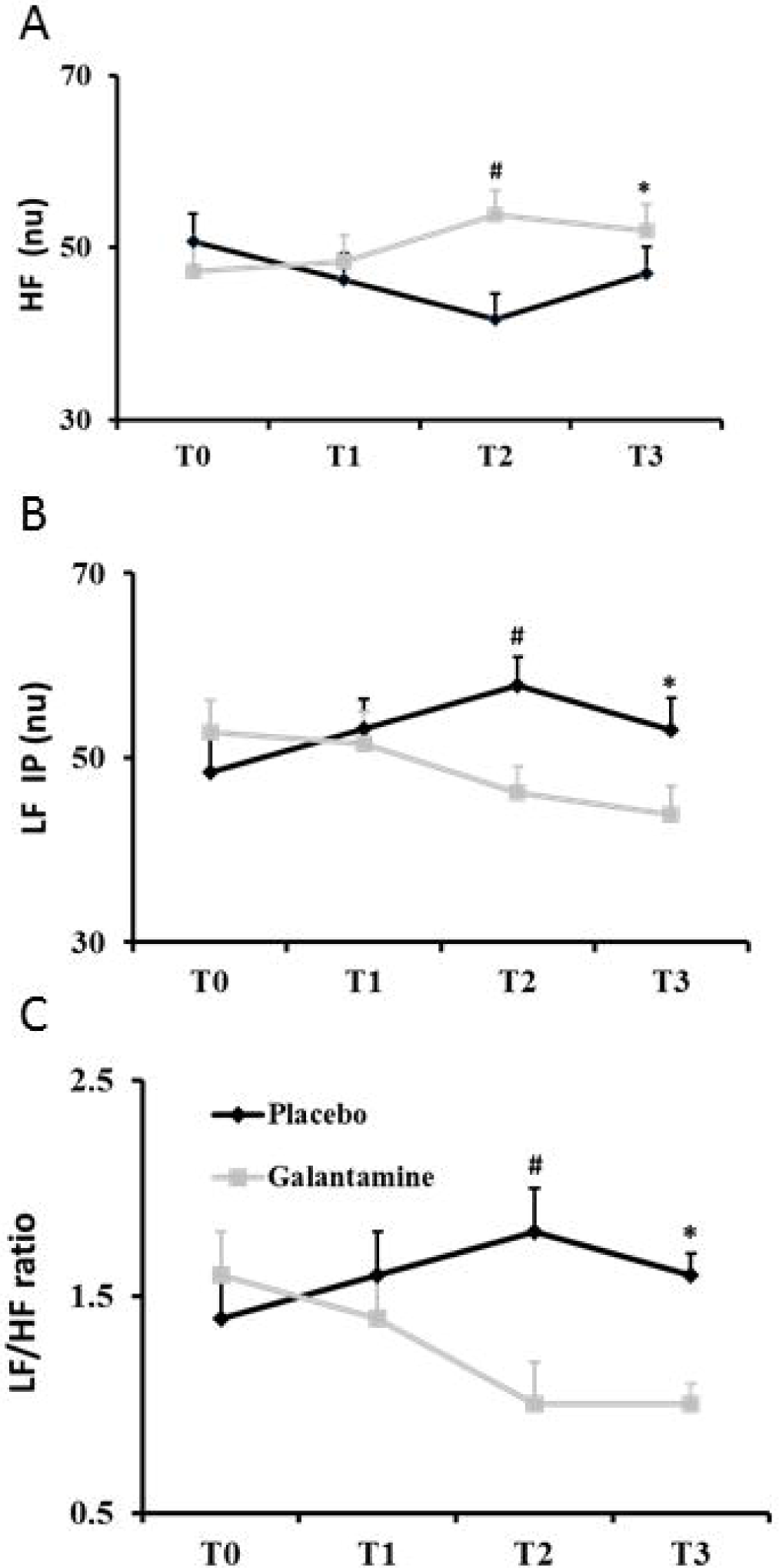
Time course of galantamine effect on heart rate variability (HRV) parameters in the frequency domain. Analysis of galantamine effects as compared to placebo was performed at randomization (T0), and after 4 (T1), 8 (T2), and 12 weeks (T3) of follow up. **(A**) Effect of galantamine on the high frequency (HF) component of HRV: ^#^P=0.007 (T2 vs T0); *P=0.04 (T3 vs T0). (**B**) Effect of galantamine on the low frequency (LF) component of HRV: ^#^P=0.009 (T2 vs T0); *P=0.005 (T3 vs T0) (**C**) Effect of galantamine on the LF/HF ratio: ^#^P=0.001 (T2 vs T0); P=0.001 (T3 vs T0).

## Discussion

To our knowledge, this is the first study to assess the effects of galantamine, a clinically approved drug for the treatment of Alzheimer’s disease on oxidative stress in patients with MetS. The results of the study demonstrate that within 12 weeks, relatively low doses galantamine significantly alleviate oxidative stress in parallel with amelioration of inflammation and insulin resistance, and beneficial alterations in autonomic regulation in MetS patients.

Although the pathogenesis of MetS remains incompletely understood, oxidative stress, chronic inflammation, and autonomic dysfunction are thought to be key constituents that can be therapeutically targeted (1, 5, 9, 10, 15). Considering the central role of oxidative stress as a driver of pathology in MetS (3, 4, 13), in this study we focused on evaluating the effects of galantamine on oxidative stress indices. Galantamine, administrated for 12 weeks significantly increased the antioxidant SOD and CAT activities and did not alter GPx activity. Lipid peroxidation determined by TBARS, another indicator of oxidative damage, was significantly decreased following galantamine treatment. Galantamine also significantly decreased plasma nitrate levels. Previously, the effects of other modalities, including mediterranean diet, melatonin, resveratrol, glycine, and vitamin E on oxidative stress in MetS have been studied (39-43). These treatments have reportedly variable effects on oxidative stress indices; some result in increased levels of SOD (39-41) while others cause a decrease in SOD-specific activity (42, 43) in parallel with affecting other indices of oxidative stress, including CAT, GPx, and malondialdehyde. However, no significant alterations in lipid and protein oxidative damage have been previously associated with these modalities. Our results, demonstrating the effects of galantamine on multiple components of oxidative and nitrosative stress in MetS patients provide novel insights extending the scope of beneficial efficacy of this clinically-approved drug.

In MetS, oxidative stress is interrelated with low-grade systemic inflammation and metabolic derangements, including insulin resistance. In this respect our results reveal galantamine anti-inflammatory effects in the cohort of 22 MetS patients in parallel with the beneficial drug effects on oxidative stress. Galantamine significantly inhibited TNF and leptin – two key mediators of the chronic inflammatory state and contributors to insulin resistance (8, 44, 45). Galantamine also increased levels of adiponectin - a molecule with anti-inflammatory properties, inversely associated with insulin resistance (8, 44, 45). The lower levels of insulin and insulin resistance in these patients indicated and strengthen the causative relationship.

In animal models, galantamine anti-inflammatory effects have been linked to activation of the vagus-nerve based inflammatory reflex (19-21, 28) and this drug has been shown to alter HRV and stimulate efferent vagus nerve activity (28, 30). Galantamine also alleviates hypertension in an animal model of lupus erythematosus (28). In addition, galantamine treatment or vagus nerve stimulation results in decreased inflammation in animal models of many diseases, including inflammatory bowel disease and arthritis with recent success in treating human conditions (19, 21, 29, 46-49). In addition to vagus nerve cholinergic signaling, sympathetic catecholaminergic outflow has been linked to modulation of inflammation (17). In many conditions, including obesity-associated disorders, aberrant inflammation coexists with autonomic dysfunction (8, 50). HRV analysis provides a non-invasive approach to examine autonomic regulation of the heart and some, although limited insights into sympatho-vagal balance (37). Autonomic dysfunction manifested by increased LF and decreased HF, alongside inflammation and oxidative stress have been reported in MetS (8, 10, 11, 15). Autonomic dysfunction has been also strongly associated with morbidity and mortality in MetS, and cardiovascular disease (6, 8, 12). Accordingly, improving HRV and lowering the LF/HF ratio have been proposed as a therapeutic approach (6, 8, 10). HRV results from the current study indicate the effect of galantamine on measures of autonomic neural modulation, as reflected by lower LF/HF ratios. This alterations were already significant at the 8 week (T2) time point, e.g. after the first 4 weeks of galantamine 16 mg/day treatment. These results suggest that galantamine effects on autonomic regulation could contribute to anti-inflammatory and insulin resistance–alleviating effects of this drug.

Treatments with AChE inhibitors, including galantamine have been shown to reduce the risk of acute myocardial infarction and death in a nationwide cohort of subjects diagnosed with Alzheimer’s dementia (34). Another study corroborates this information (51). A recent meta-analysis-based study highlights that galantamine and other AChE inhibitor treatments are associated with lower risk of CV events including stroke, acute coronary syndrome, and cardiovascular mortality (52).

It should be noted that the beneficial effects of galantamine in MetS patients were observed using relatively low doses. The highest dose used (16 mg/day) in this study is less than the highest dose - 24 mg/day, approved for the treatment of AD patients with typically much lower BMIs. The study subjects tolerated well this dosage and none of them reported adverse effects. While galantamine significantly altered autonomic modulation (based on HRV analysis), no significant alterations were observed in heart rate and office BP. These findings are in line with a previous report in Alzheimer’s patients treated with galantamine (53). The absence of adverse effects on cardiovascular and metabolic parameters indicate a good safety profile of galantamine treatment at the doses used.

In conclusion, our results demonstrate that treatment with relatively low doses galantamine, a drug in clinical use for the symptomatic treatment of Alzheimer’s disease, alleviates oxidative stress in patients with MetS. They also confirm the beneficial drug effects on inflammation, insulin resistance and autonomic modulation in these patients. These results, alongside previously published findings (31) considerably strengthen the rationale for further studies with galantamine for therapeutic benefit in the growing at-risk population of people with MetS.

## Data Availability

The datasets used and/or analysed during the current study are available from the corresponding authors on reasonable request.

## Ethics approval and consent to participate

The study protocol was reviewed and approved by the Institutional Review Committee and the Human Subject Protection Committee of the Heart Institute (InCor) and the Clinic Hospital (number 11672/555738), University of São Paulo. The study was conducted in accordance with World Medical Association International Code of Medical Ethics (Declaration of Helsinki, 1964; revised in 2008). The study is registered at www.clinicaltrials.gov with the number NCT02283242 and the following name: “Galantamine effects in patients with metabolic syndrome (GALANTA-MS)”: https://clinicaltrials.gov/ct2/show/NCT02283242?term=galantamine+metabolic+syndrome&rank=1

## Consent for publication

All study participants provided written informed consent for publication as part of their consent for participation in the research study.

## Availability of data and materials

The datasets used and/or analysed during the current study are available from the corresponding author on reasonable request.

## Authors’ contributions

FMCC and VAP designed the study. CTS, KYK, TLM, AAA, KDA acquired the data. CTS and LCM performed the statistical analyses. FMCC, CTS, KDA, MCI and LCM interpreted the data. CTS wrote the first draft of the report with inputs from FMCC and LCM. FMCC, CTS, LCM, KYK, KDA, MCI, TLM VAP, DPB, and KJT provided comments, participated in additional discussions and revised the paper. VAP, DPB, KJT, and FMCC edited the text and finalized the manuscript. All authors approved the final version.

## Acknowledgements

This study was supported by Fundação de Amparo a Pesquisa do Estado de São Paulo (FAPESP), Conselho Nacional de Desenvolvimento Científico e Tecnológico (CNPq) and the NIH. The funders of this study had no role in study design, data collection, data analysis, data interpretation or manuscript writing. The corresponding author had full access to all the data of the study and had the final responsibility for the manuscript submission.

